# A multi-database based ceRNA regulatory network for gastric cancer prognosis

**DOI:** 10.1101/2023.08.01.23293496

**Authors:** Long-Kuan Yin, Qian Li

## Abstract

**Objective:** Circular RNA(circRNA) is a kind of endogenous non-coding RNA, which may be related to the occurrence and development of cancer. Based on the GEO database, this paper constructs a circRNA as a competitive endogenous RNAs(ceRNAs) that binds with microRNAs (miRNAs) to affect and regulate the expression of target genes. The ceRNA regulatory network based on circRNA-miRNA-mRNA model plays an important role in tumor prognosis and treatment. This paper explores the mechanism of circRNA-related ceRNA regulatory network in gastric cancer.

**Method:** CircRNA, miRNA and mRNA data sets related to gastric cancer were downloaded from The Gene Expression Omnibus (GEO), and the limma package of R software (R 4.2.1 version) was used to identify the differences between gastric cancer tissues and adjacent normal tissues of gastric cancer. DEcircRNA, DEmiRNAs, and DEmRNAs. Based on circBase database, we explored the interactions among circRNA, miRNA and mRNA, and constructed the circRNA-miRNA-mRNA ceRNA network by using Cytoscape_v3.8.0. Then KEGG, GO and survival analysis of ceRNA-related genes were performed. Then, the prognostic data of gastric cancer were extracted from the TCGA database to construct the prognostic subnetwork of gastric cancer.

**Results:** KEGG analysis of ceRNA’s mRNA showed that the pathway was mainly enriched in IL-17 signaling pathway, TNF signaling pathway and so on, which affected the prognosis of gastric cancer. hsa_circ_0055521/hsa-miR-204-5p/FAP, (hsa_circ_0005051, hsa_circ_0007613, hsa_circ_0045602, hsa_circ_0034398, hsa_circ_0006089) /hsa-miR-32-3p/FNDC1 were the ceRNA networks related to the prognosis of gastric cancer collaterals.

**Conclusion:** This study found that IL-17 signaling pathway and TNF signaling pathway may affect the occurrence, development and prognosis of gastric cancer. And hsa_circ_0055521/hsa-miR-204-5p/FAP, (hsa_circ_0005051, hsa_circ_0007613, hsa_circ_0045602, hsa_circ_0034398, hsa_circ_0006089) /hsa-miR-32-3p/FNDC1 two circrNa-based stomachs The cancer ceRNA prognostic network is a new prognostic related ceRNA network for gastric cancer. FNDC1 and FAP may be potential therapeutic targets for gastric cancer.

## 1. Introduction

Gastric cancer (GC), as a common global cause of cancer death [1-3], is most frequently found in East Asia, such as South Korea, China and Japan[4,5] and so on. The early clinical symptoms of gastric cancer lack specificity [6-8], which leads to the late diagnosis of most gastric cancer patients[9]. Although the diagnosis and treatment strategies for gastric cancer have gradually increased in recent decades, the treatment effectiveness of advanced gastric cancer is poor, and the prognosis of patients remains poor[10-12]. The complexity and multi-factor nature of pathogenic factors, occurrence, development and metastasis mechanism of gastric cancer make the treatment of gastric cancer more difficult and complicated. Patients with advanced gastric cancer have a lower 5-year survival rate[13]. Therefore, there is an urgent need to find more therapeutic targets to improve diagnosis and treatment measures to improve the survival and prognosis of patients.

In recent years, with the development of gene technology, we have found that there are very rich regulatory modes of various Rnas in cells, and ceRNA regulation is one of the more studied regulatory modes. ceRNA refers to a new mechanism of RNA interaction in which mRNA, lncRNA, circRNA, etc. regulate each other at the post-transcriptional level through competition and sharing of miRNA[14].

CircRNAs are a class of covalently closed single-stranded RNA molecules that exist widely in eukaryotic cells. circRNA is a class of regulatory molecules with various functions, which can play functions such as regulating gene expression, binding miRNA or protein, and translating into protein, etc. CircRNA may be of great significance for cell proliferation, cell differentiation, individual growth and development, and may be related to the occurrence and development of cancer [15,16]. Based on the GEO database, this paper constructs a circRNA as a competitive endogenous RNAs(ceRNAs) that binds with microRNAs (miRNAs) to affect and regulate the expression of target genes. The ceRNA regulatory network based on circRNA-miRNA-mRNA model plays an important role in tumor prognosis and treatment. This paper explores the mechanism of circRNA-related ceRNA regulatory network in gastric cancer.

CircRNA, miRNA and mRNA data sets related to gastric cancer were downloaded from The Gene Expression Omnibus (GEO)[17] and limma package of R software (R 4.2.1 version) was used to identify the relationship between gastric cancer tissue and adjacent normal tissue. Differentially expressed circRNA (DEcircRNA), differentially expressed miRNAs (DEmiRNAs) and differentially expressed mRNAs (DEmRNAs). We analyzed the relationship between Rnas based on circBase database, and constructed a ceRNA network using circRNA-miRNA-mRNA template by using Cytoscape_v3.8.0. Then, the target genes of ceRNA were analyzed by KEGG[18], GO[19]and survival analysis. The mRNA expression and clinical data related to gastric Cancer were extracted from The Cancer Genome Atlas[20] (TCGA) database, and the survival data of gastric cancer were extracted. Then, combined with the relevant gastric cancer data of TCGAand GEO databases, we explored the potential relationship between circRNA, miRNA and mRNA and the prognosis of gastric cancer, searched for new potential ceRNA regulatory networks, and searched for potential therapeutic targets through this network.

## 2. Materials and methods

2.1 Data download and processing

Downloaded from GEO database (https://www.ncbi.nlm.nih.gov/geo/) gastric cancer related GSE83521 circRNA dataset, the miRNA GSE93415 data sets, mRNA GSE118916 data sets.

2.2 Difference analysis was conducted on circRNA, miRNA and mRNA data respectively.

Firstly, the difference analysis of circRNA, miRNA and mRNA was carried out respectively, and the heat map analysis of circRNA, miRNA and mRNA with significant differences was carried out respectively.

2.3 CircRNA, miRNA and mRNA of ceRNA network were extracted and the ceRNA network diagram was constructed.

First, the structure diagram of circRNA and the set MRE of reaction elements were found through circBase database. The intersection of mirna.diff and MRE is made by Venn diagram. The target file was obtained by predicting the target gene of the co-expressed micRNA, and the target and mRNA. diff files were processed by venn using R software (R 4.2.1 version) to search for the co-expressed mRNA. Then prepare MRE, mrnas.diff, target, mirna.diff, circ.diff files respectively, and use Perl software to process each file. The files ceRNA.network, ceRNA.node, ceRNA.circList, ceRNA.mirnaList, and cerna.mrnalist are obtained respectively. Finally, the ceRNA network diagram is established using Cytoscape 3.8.0.

2.4 The expression differences of circRNA, miRNA and mRNA between gastric cancer tissues and adjacent normal tissues were analyzed.

Heat map analysis and boxplot analysis of ceRNA’s RNA expression between tumor tissue and normal tissue.

2.5 mRNA in ceRNA network was analyzed by GO and KEGG.

mRNA in ceRNA network was analyzed by GO and KEGG to find the possible signaling pathway.

2.6 Gastric cancer expression data and clinical data were extracted from the TCGA database for survival analysis.

Will be extracted from TCGA database (https://portal.gdc.cancer.gov/) of gastric cancer expression data sorting for expressing, extract gastric cancer clinical data. The gastric cancer expression profile was combined with clinical data for survival analysis.

2.7 Gastric cancer data of TCGA and GEO databases were merged.

The expression index and clinical data of gastric cancer extracted from TCGA database and RNA from ceRNA network from GEO database were combined to obtain the survival data of ceRNA network. We then performed ceRNA survival analysis.

2.8 Construct ceRNA network diagram related to prognosis of gastric cancer.

Based on the above data, we constructed a prognostic correlation network map for gastric cancer.

2.9 Protein immunohistochemical analysis was performed by HPA. Protein immunohistochemical analysis of prognostic ceRNA network mRNA was performed using HPA database.

2.10 Statistical analysis. All statistical analyses were performed using R software. *P* < 0.05 was considered statistically significant.

## 3. Results

3.1 Differences of circRNA, miRNA and mRNA were analyzed respectively.

We found 24 significantly different circRNAs, 91 significantly different miRNAs, and 114 significantly different mRNAs through difference analysis. Through heat map analysis, we found 24 significantly different circRNAs with low expression in normal tissues and almost high expression in tumor tissues (Figure 1, A). The expression of 38 miRNAs was high in normal tissues and low in tumor tissues. Fifty-three miRNAs were expressed in normal tissues and highly expressed in tumor tissues (Figure 1, B). 82 mRNA were highly expressed in normal tissues and low expressed in tumor tissues. 32 mRNAs were low expressed in normal tissues and high expressed in tumor tissues (Figure 1,C).

**Figure 1.**
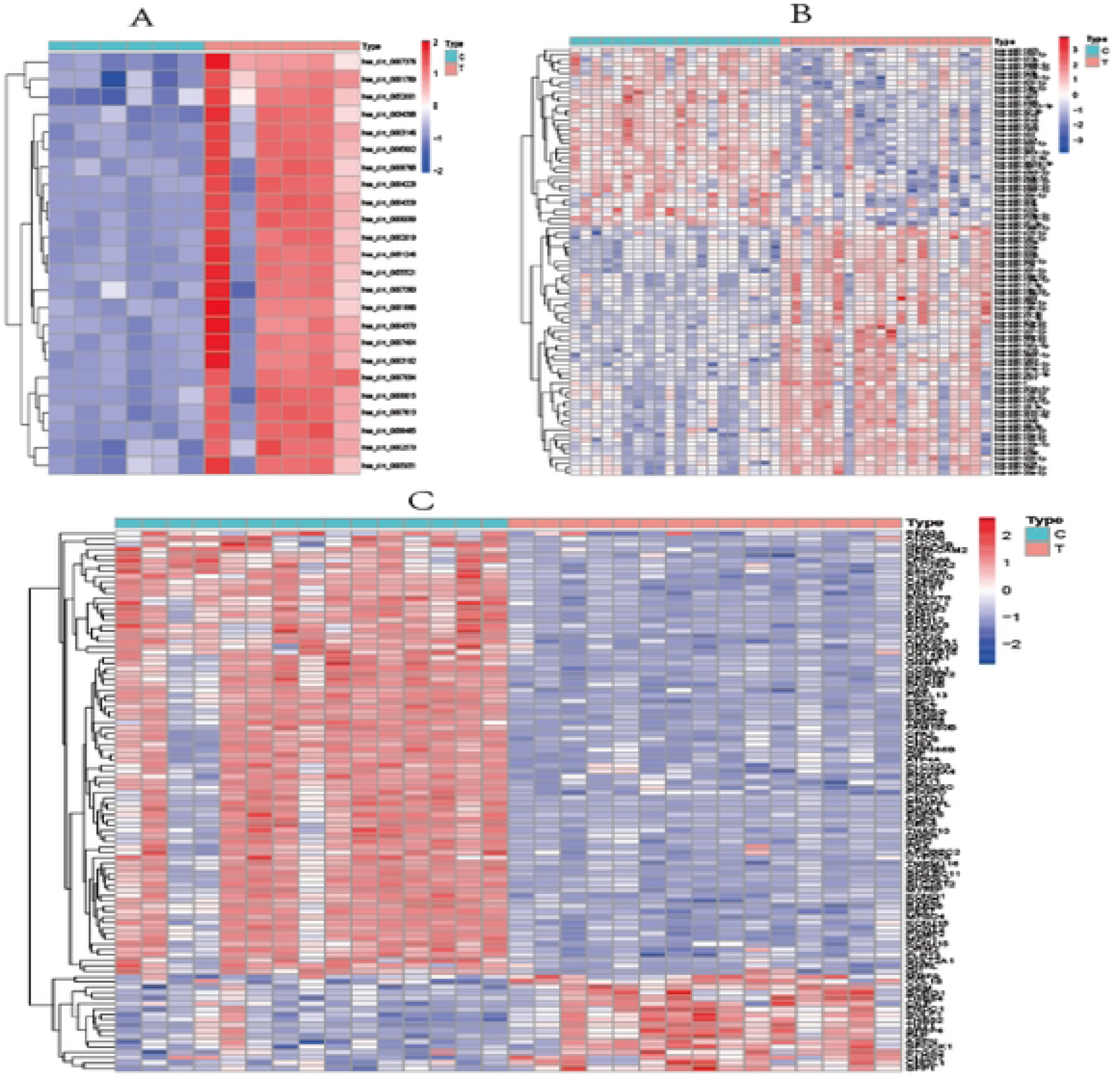
A: Heat maps of 24 circRNAs with significant diITcrcnccs; B: Heat maps of 91 miRNAs with significant differences; C: Heal maps of 114 significantly different mRNAs.

3.2 The circRNA structure diagram (Figure 2.1-2.2) and the set MRE of reaction elements were searched through circBase database. miRNA.diff and MRE were intersected by Venn diagram (Figure 2.3, A). The target file was obtained by predicting the target gene of the co-expressed micRNA, and the target and mRNA. diff files were processed by venn using R software (R 4.2.1 version) to find the co-expressed mRNA (Figure 2.3, B). Then prepare MRE, mRNAs.diff, target, miRNA.diff, circRNA.diff files respectively, and use Perl software to process each file. The files ceRNA.network, ceRNA.node, ceRNA.circList, ceRNA.mirnaList, and ceRNA.mrnalist are obtained respectively. Finally, the ceRNA network diagram is constructed with Cytoscape 3.8.0. We found that 13 circRNAs, 11 miRNAs, and 15 mRNAs formed the ceRNA network nodes. The ceRNA network diagram (Figure 2.3) was constructed.

**Figure 2.1.**
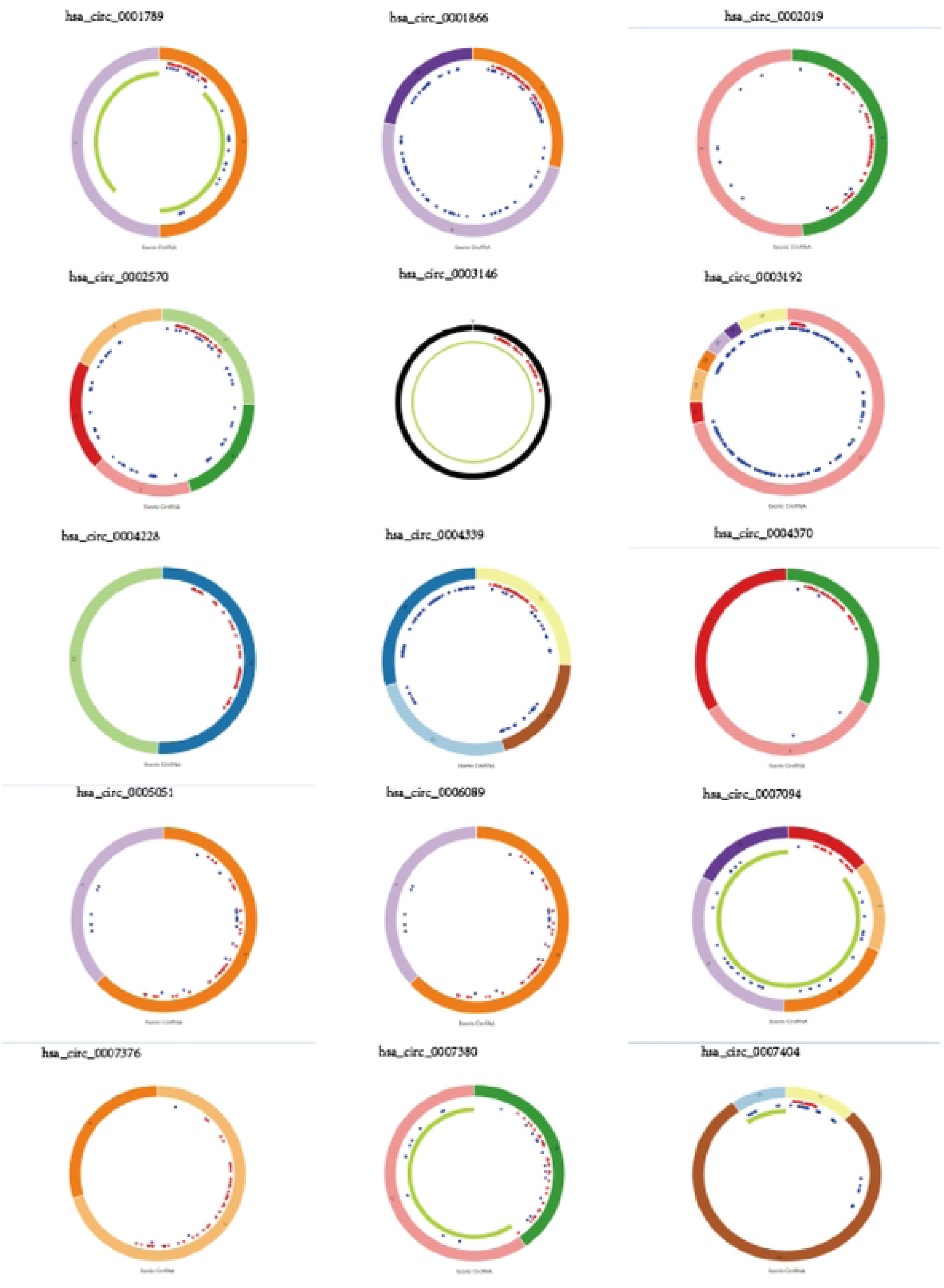
Structure diagram of part circRNA;

**Figure 2.2.**
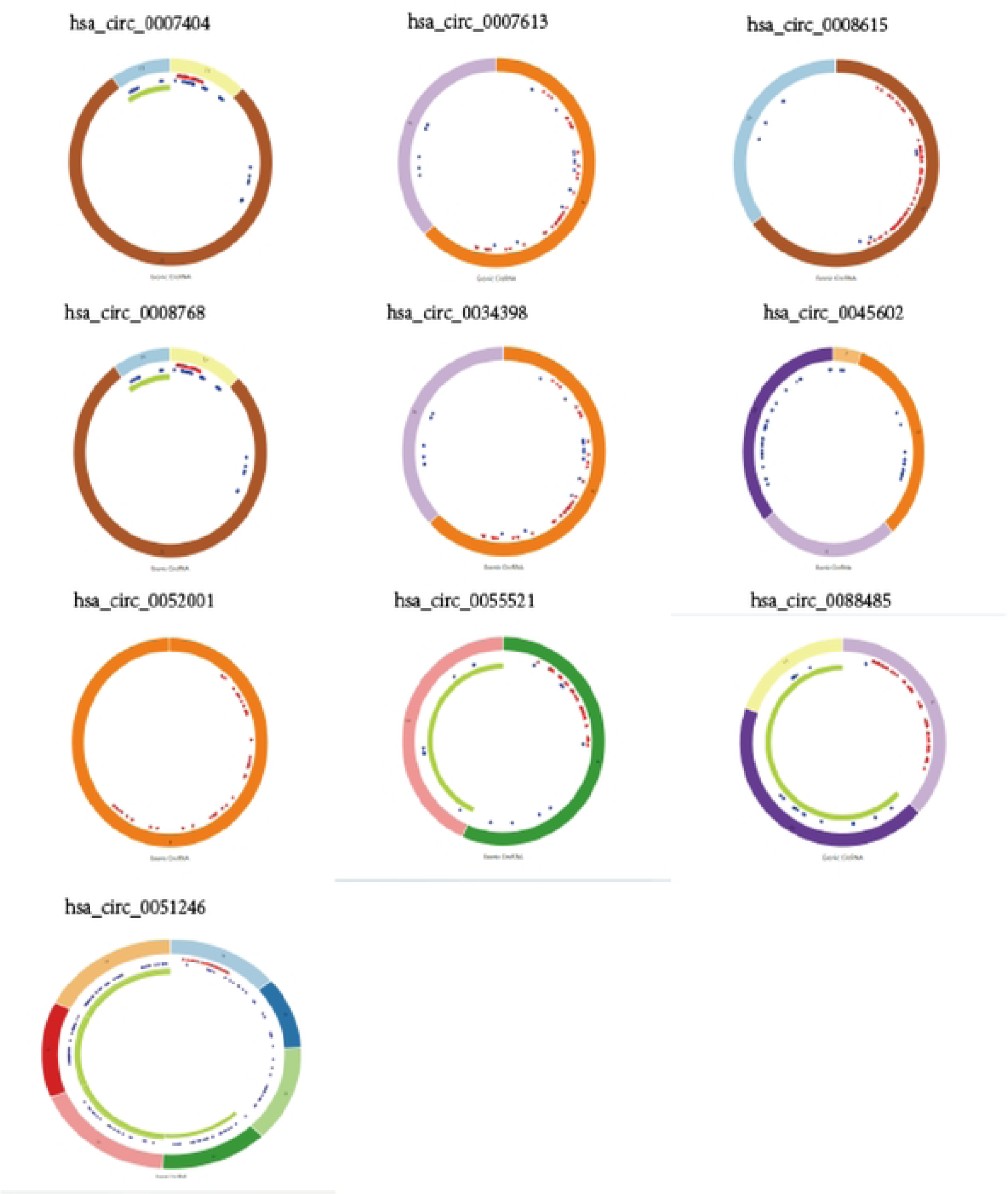
Structure diagram of partial circRNA;

**Figure 2.3.**
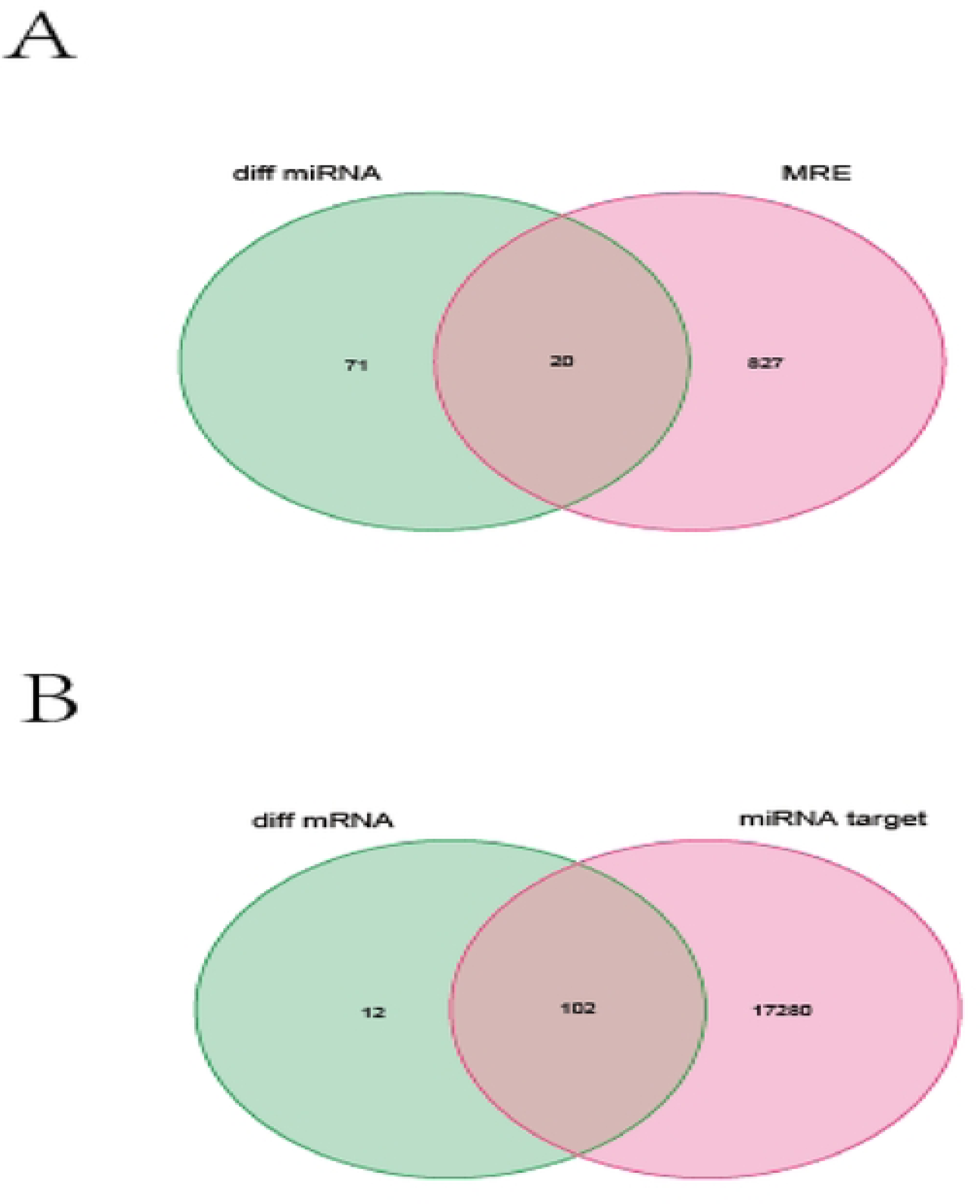
A: miRNA.diITand MRE are intersected by Venn diagram; B: target and mRNA.diIT looks for mRNA that is co-expressed.

3.3 Through heat map analysis of circRNA, miRNA and mRNA that constitute ceRNA network, it can be found that 13 circRNAs have low expression in normal samples and high expression in tumor samples. The expression of 11 miRNAs was high in normal samples and low in tumor samples. The expression of 15 mRNA was low in normal samples and high in tumor samples. A ceRNA network diagram is constructed (Figure 3).

**Figure 3.**
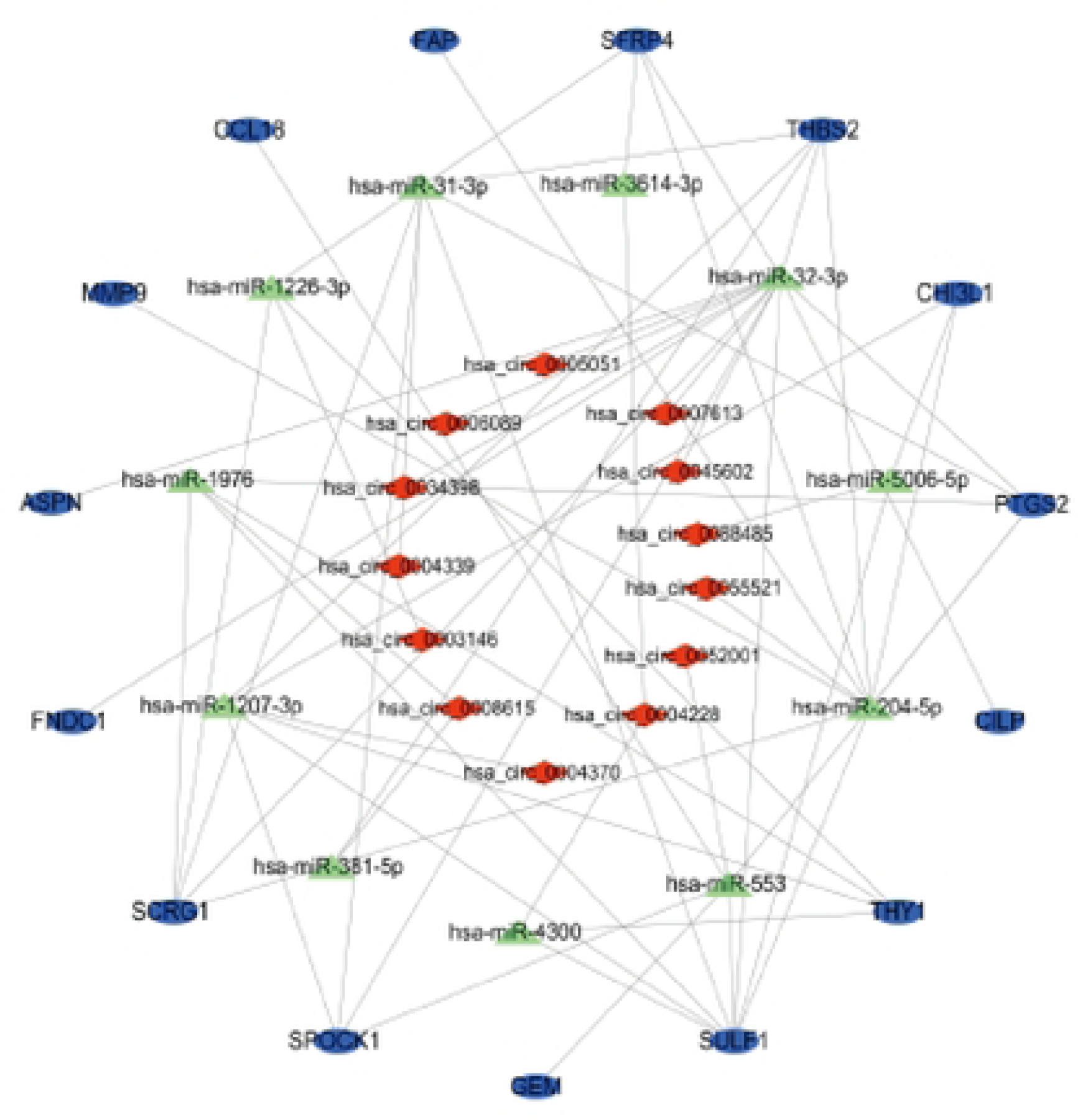
ccRNA network diagram built via Cytoscapc 3.8.0.

3.4 Through the analysis of heat maps (Figure 4.1A, B, Figure 4.2) and box plots (Figure 4.3-4.4) of circRNA, miRNA and mRNA constituting ceRNA network, it can be found that there are differences in circRNA, miRNA and mRNA between tumor tissues and normal tissues.

**Figure 4.1.**
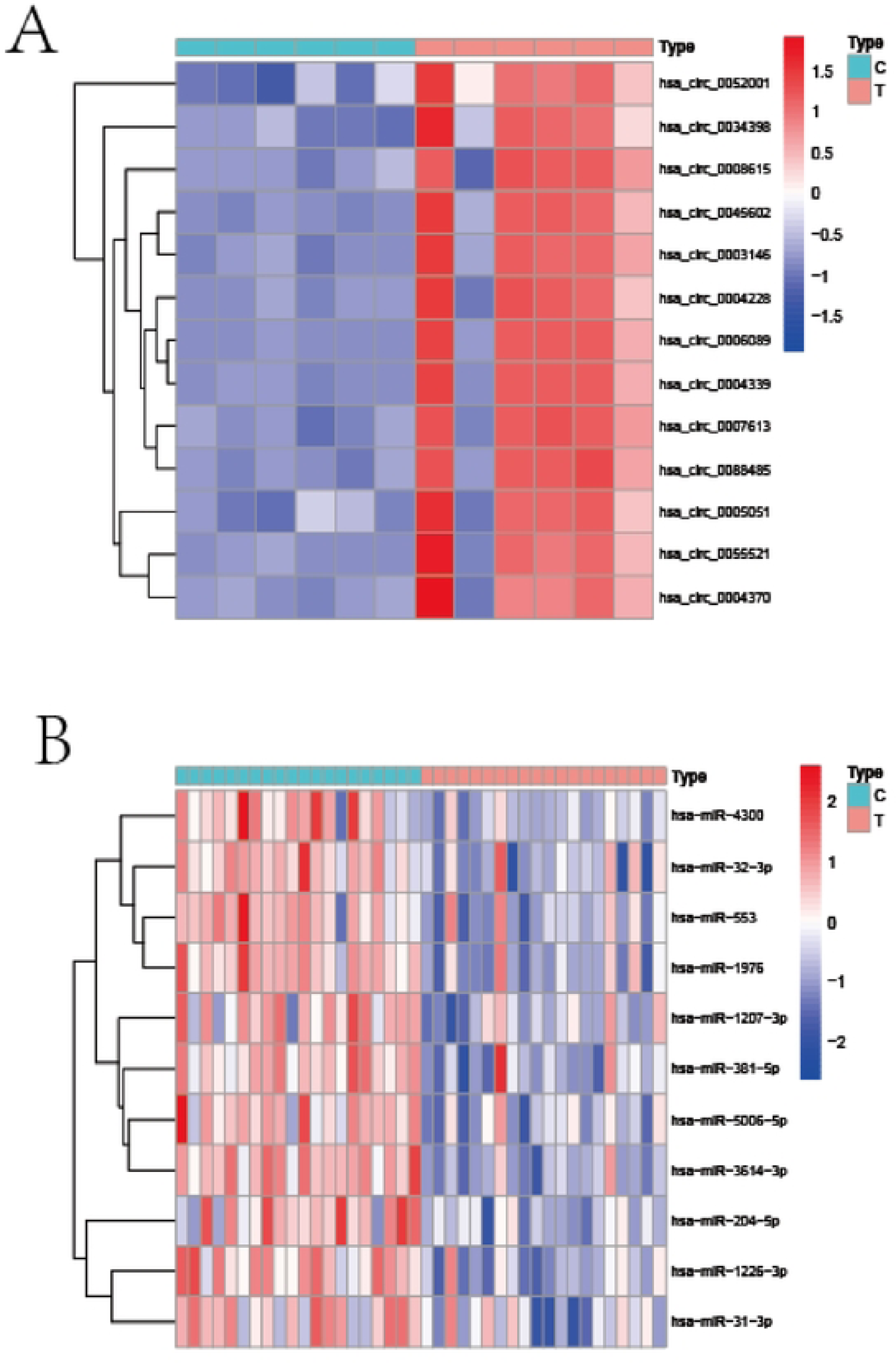
A: circRNA heat map of ccRNA network; B: miRNA heat map of ccRNA network.

**Figure 4.2.**
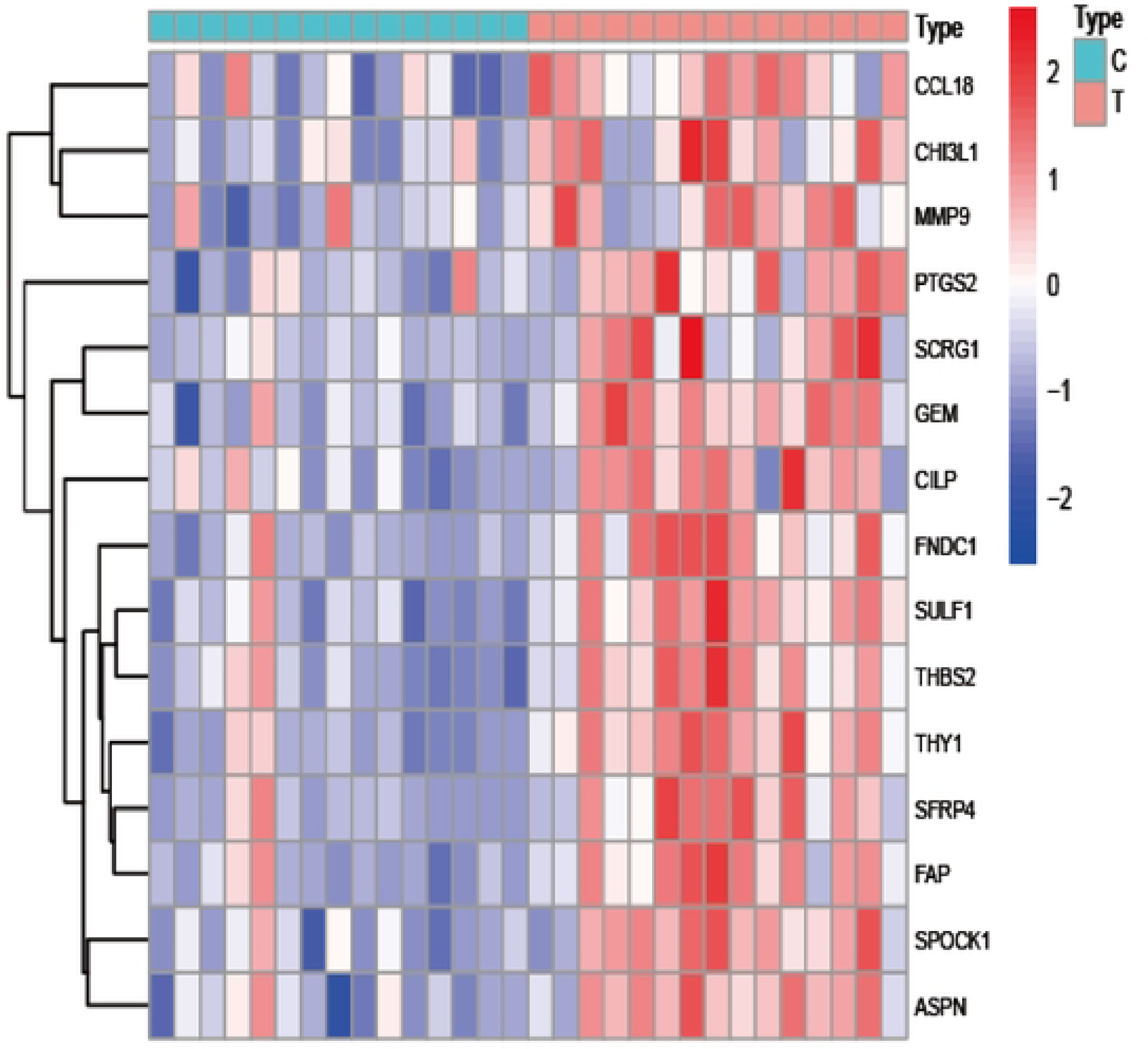
mRNA heat map of ceRNA network.

**Figure 4.3.**
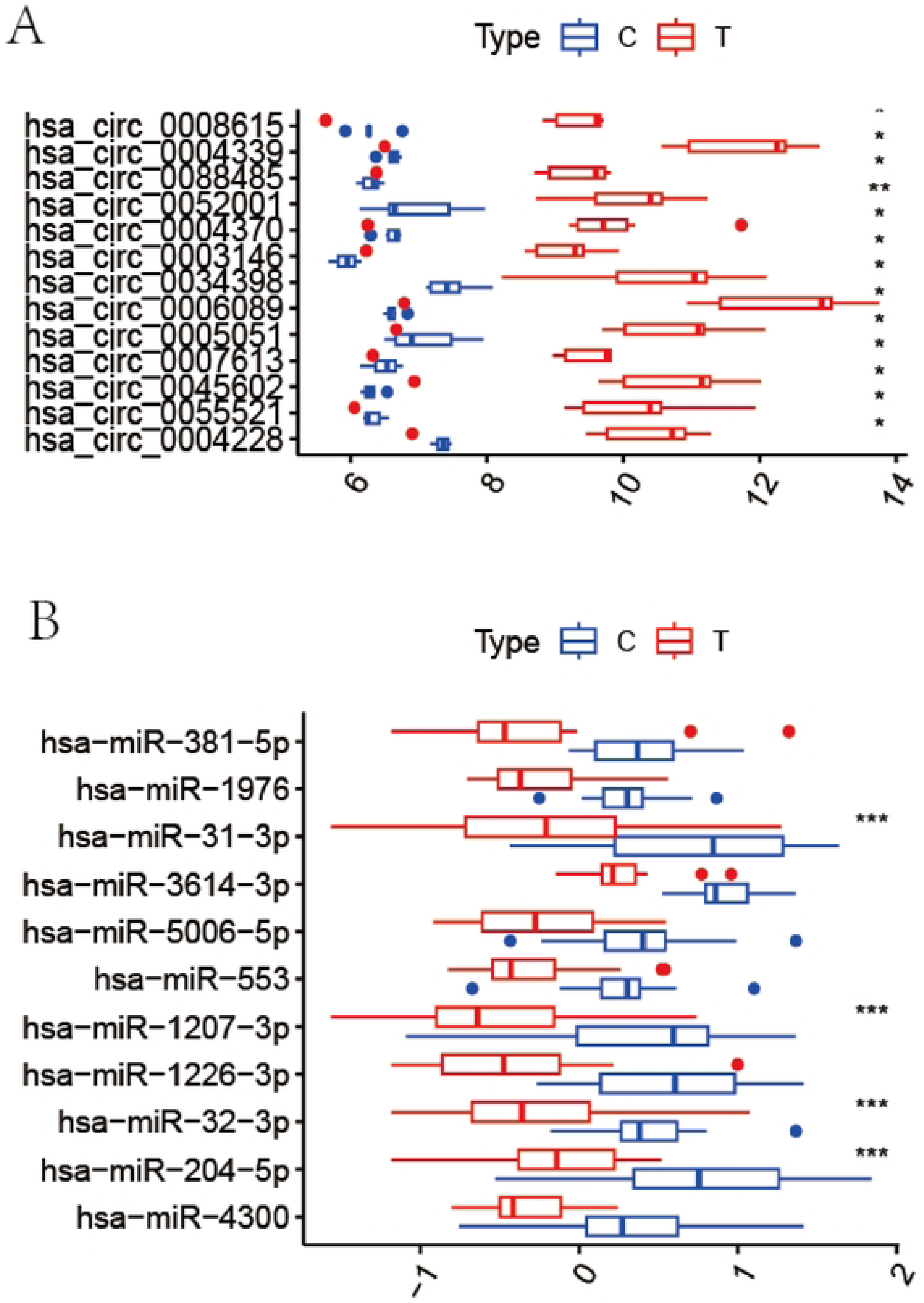
A: circRNA box diagram of ccRNA network; B: miRNA box diagram of ccRNA network.

**Figure 4.4.**
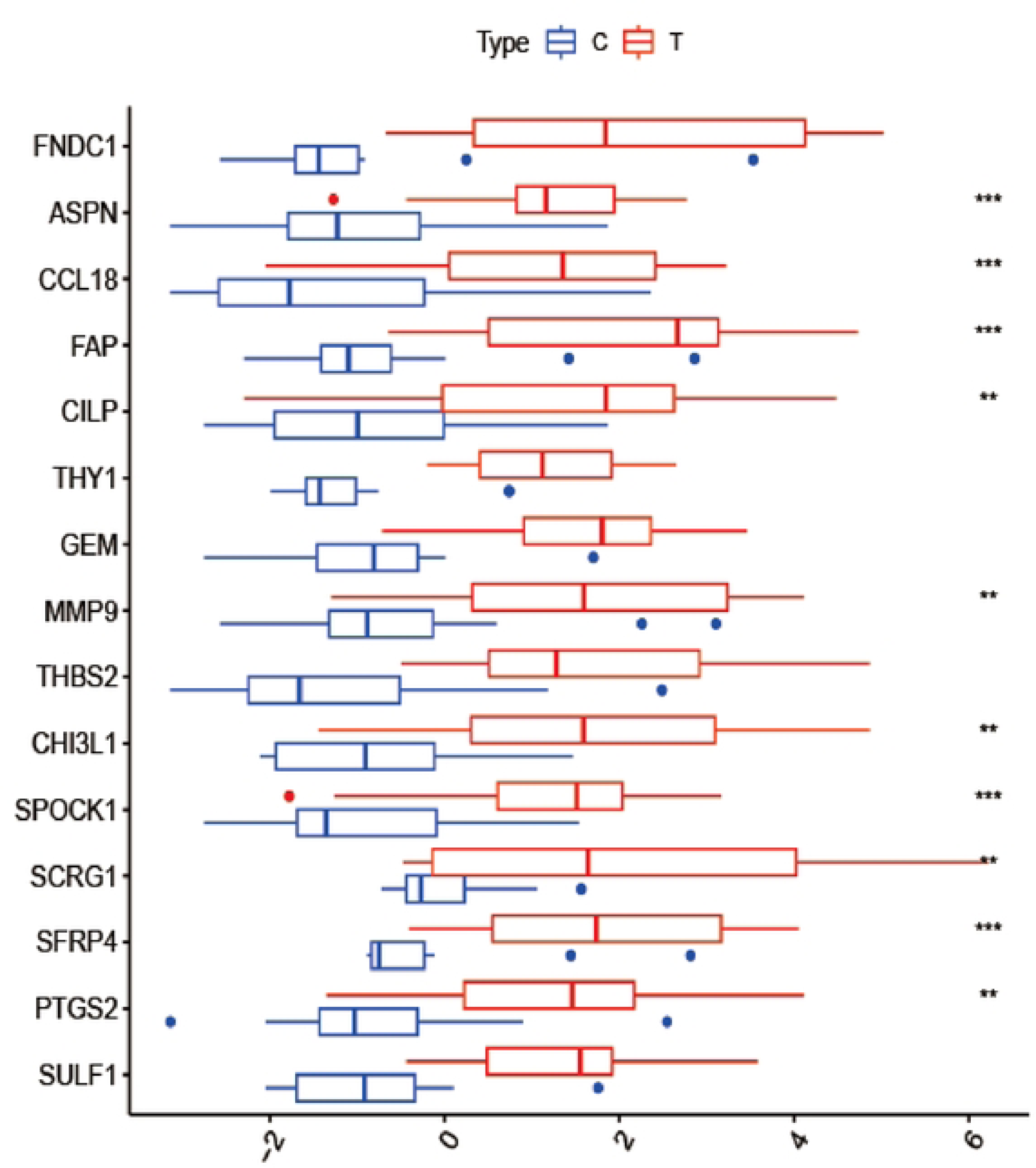
mRNA box diagram of ceRNA network.

3.5 GO (Figure 5.1-5.3,table 1-3) and KEGG (Figure 5.4,table 4) analyses were performed on mRNA constituting the ceRNA network. In this study, it was found that the main pathways were enriched in IL-17 signaling pathway and TNF signaling pathway.

**Figure 5.1.**
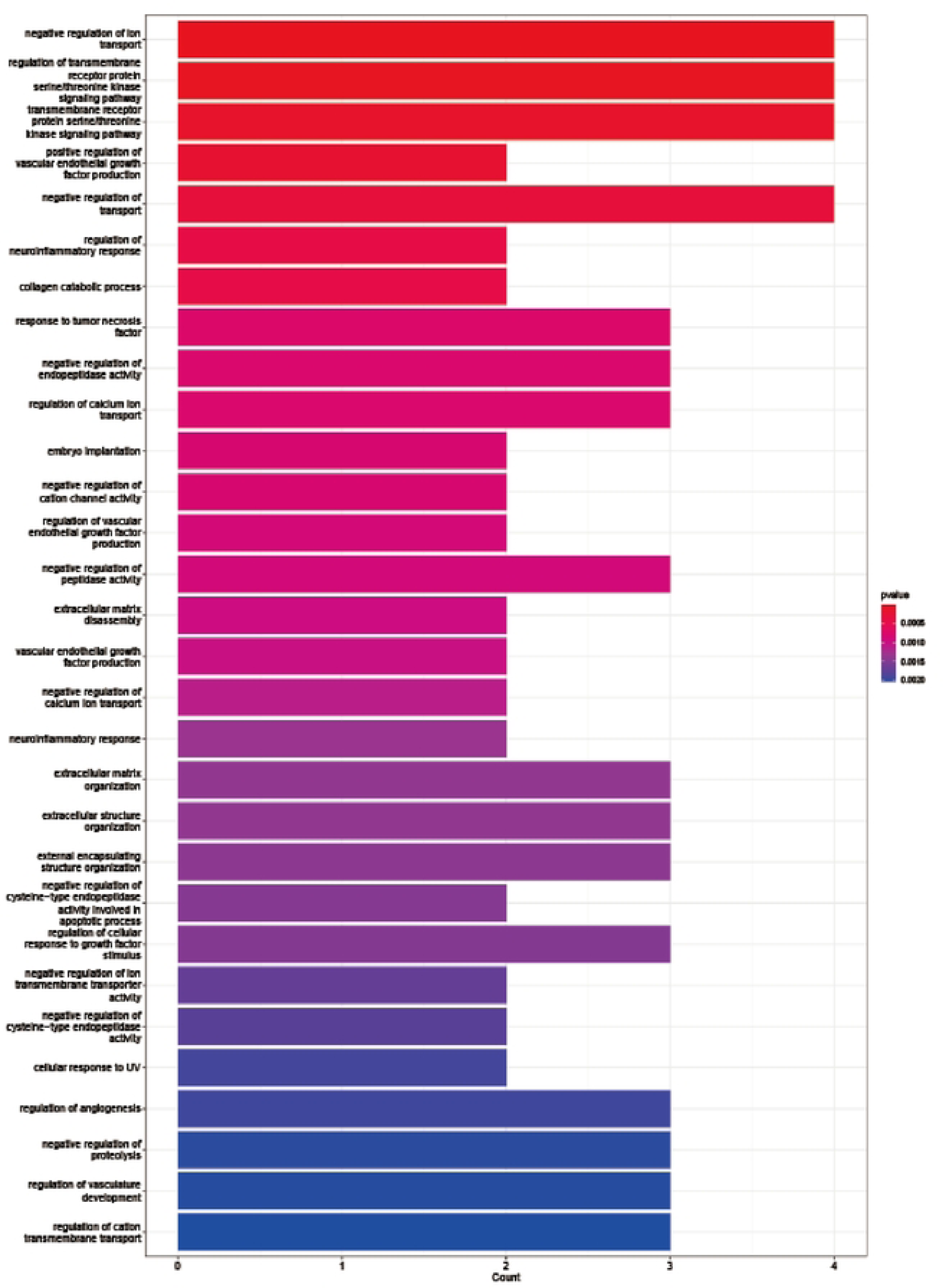
BPenrichment map for GO analysis ofmRNA constituting ceRNA network.

**Figure 5.2.**
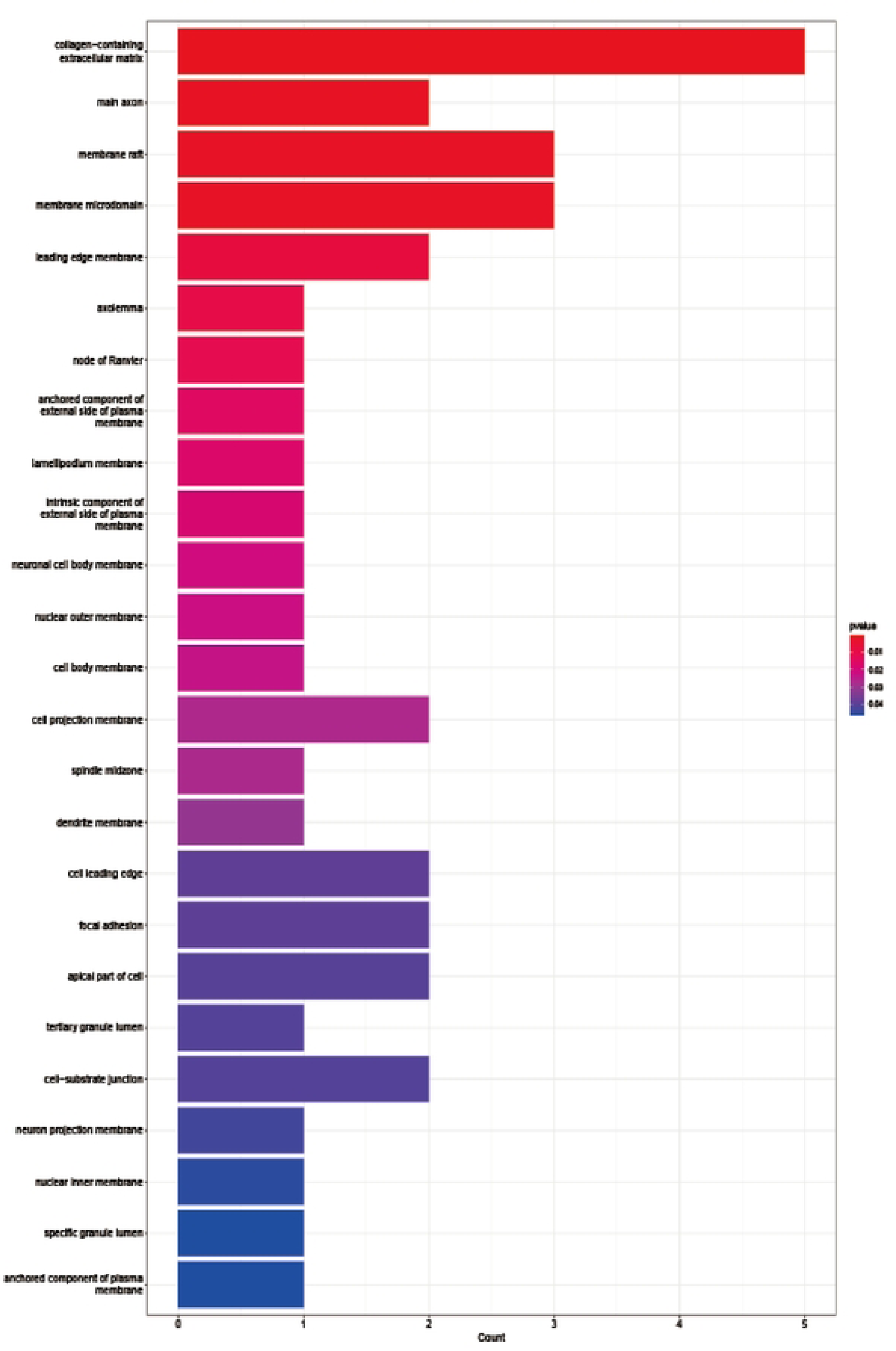
CC enrichment map for GO analysis of mRNA constituting ceRNA network.

**Figure 5.3.**
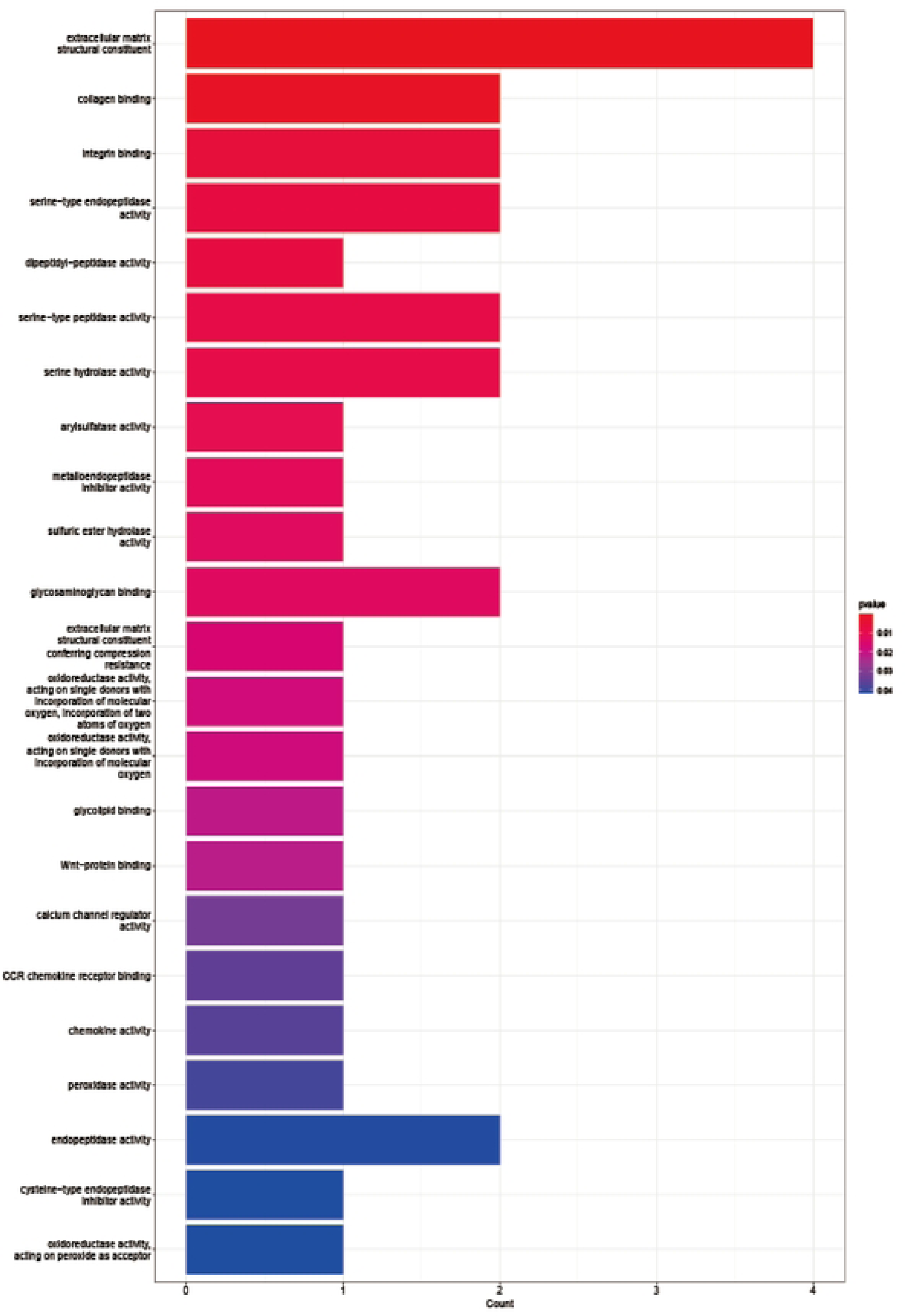
MF enrichment diagram for GO analysis of mRNA constituting ceRNA network.

**Figure 5.4.**
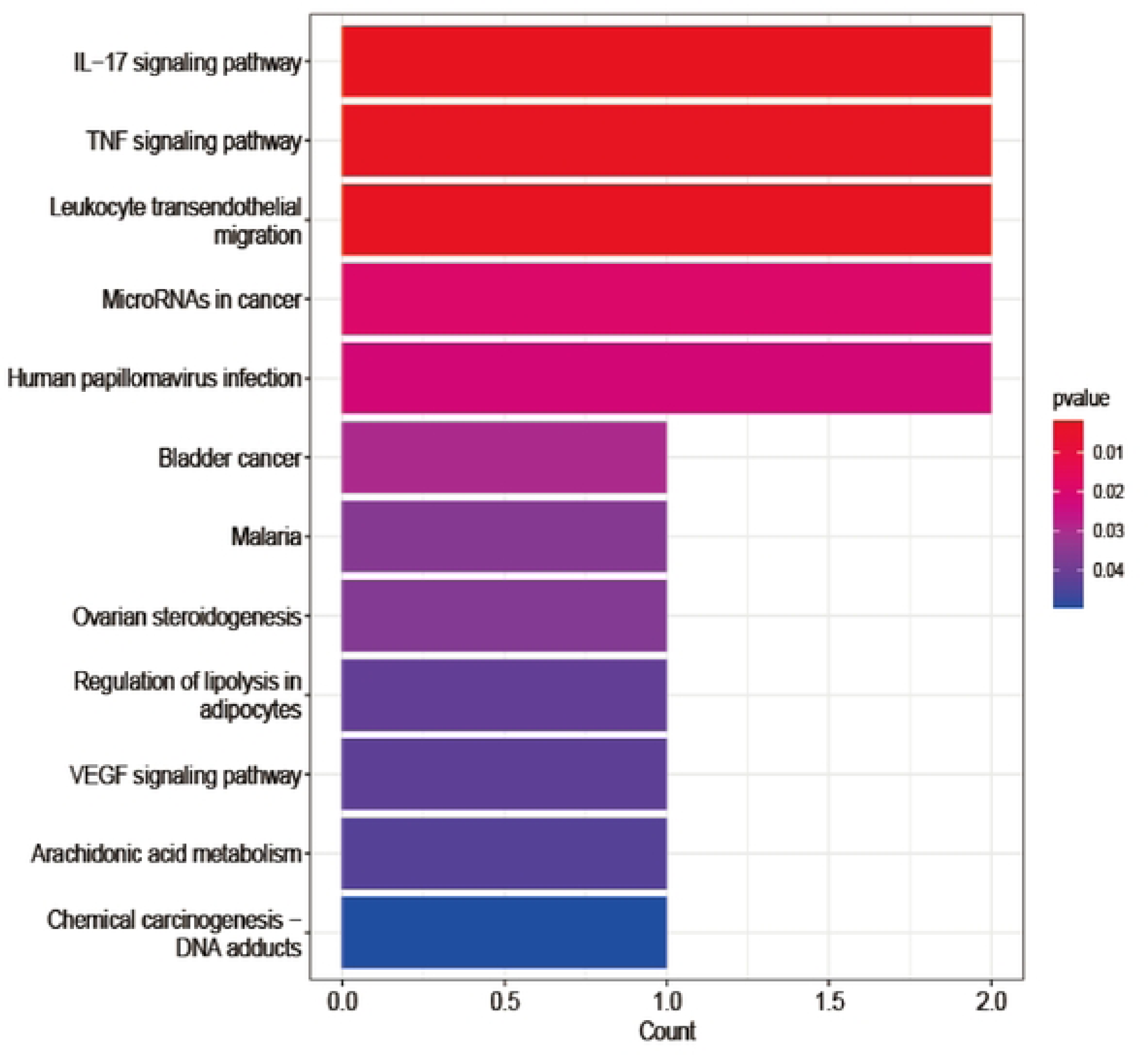
Pathway map for KEGG enrichment analysis ofmRNA constituting ccRNA network.

**Table 1.** The top 20 BP results of GO analysis.

| ID | Description | pvalue | geneID |
| --- | --- | --- | --- |
|  | negative regulation of ion transport | 4.17E-06 | PTGS2/SFRP4/MMP9/GEM |
|  | regulation of transmembrane receptor protein serine/threonine kinase signaling pathway | 3.56E-05 | SULF1/SFRP4/CILP/ASPN |
|  | transmembrane receptor protein serine/threonine kinase signaling pathway | 0.000123768 | SULF1/SFRP4/CILP/ASPN |
|  | positive regulation of vascular endothelial growth factor production | 0.000206688 | SULF1/PTGS2 |
|  | negative regulation of transport | 0.000301177 | PTGS2/SFRP4/MMP9/GEM |
|  | regulation of neuroinflammatory response | 0.000415322 | PTGS2/MMP9 |
|  | collagen catabolic process | 0.000435902 | MMP9/FAP |
|  | response to tumor necrosis factor | 0.000750016 | PTGS2/CHI3L1/CCL18 |
|  | negative regulation of endopeptidase activity | 0.000767632 | PTGS2/SPOCK1/MMP9 |
|  | regulation of calcium ion transport | 0.000767632 | PTGS2/GEM/THY1 |
|  | embryo implantation | 0.000802875 | PTGS2/MMP9 |
|  | negative regulation of cation channel activity | 0.000802875 | MMP9/GEM |
|  | regulation of vascular endothelial growth factor production | 0.000859994 | SULF1/PTGS2 |
|  | negative regulation of peptidase activity | 0.000869263 | PTGS2/SPOCK1/MMP9 |
|  | extracellular matrix disassembly | 0.000949255 | MMP9/FAP |
|  | vascular endothelial growth factor production | 0.000979961 | SULF1/PTGS2 |
|  | negative regulation of calcium ion transport | 0.001107533 | PTGS2/GEM |
|  | neuroinflammatory response | 0.001313061 | PTGS2/MMP9 |
|  | extracellular matrix organization | 0.001373366 | SULF1/MMP9/FAP |
|  | extracellular structure organization | 0.001386263 | SULF1/MMP9/FAP |

**Table 2.** Thetop20CCresultsofGO analysis.

The top 20 CC results of GO analysis.
| ID | Description | pvalue | geneID |
| --- | --- | --- | --- |
|  | collagen-containing<br>extracellular matrix | 1.23E-05 | SULF1/THBS2/MMP9/<br>CILP/ASPN |
|  | main axon | 0.000975247 | SPOCK1/THY1 |
|  | membrane raft | 0.001789774 | SULF1/PTGS2/THY1 |
|  | membrane microdomain | 0.001805514 | SULF1/PTGS2/THY1 |
|  | leading edge membrane | 0.007715433 | THY1/FAP |
|  | axolemma | 0.009909463 | THY1 |
|  | node of Ranvier | 0.011425836 | SPOCK1 |
|  | anchored component of external<br>side of plasma membrane | 0.01520729 | THY1 |
|  | lamellipodium membrane | 0.016716088 | FAP |
|  | intrinsic component of external<br>side of plasma membrane | 0.018222729 | THY1 |
|  | neuronal cell body membrane | 0.021229548 | THY1 |
|  | nuclear outer membrane | 0.021979909 | PTGS2 |
|  | cell body membrane | 0.02347902 | THY1 |
|  | cell projection membrane | 0.027010044 | THY1/FAP |
|  | spindle midzone | 0.027217419 | GEM |
|  | dendrite membrane | 0.030942458 | THY1 |
|  | cell leading edge | 0.039342466 | THY1/FAP |
|  | focal adhesion | 0.039860127 | THY1/FAP |
|  | apical part of cell | 0.040728808 | THY1/FAP |
|  | tertiary granule lumen | 0.04130 | MMP9 |

**Table 3.**
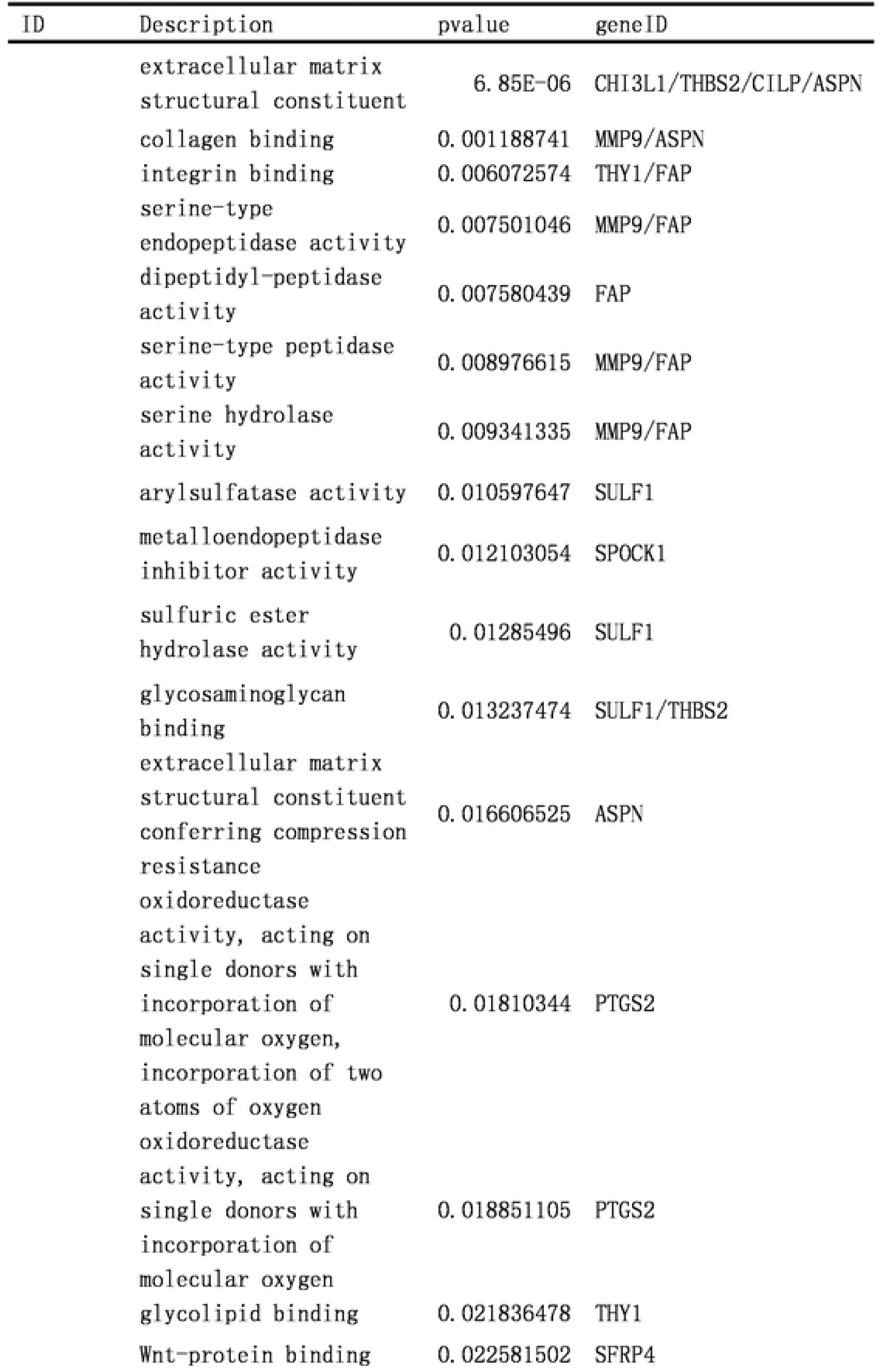
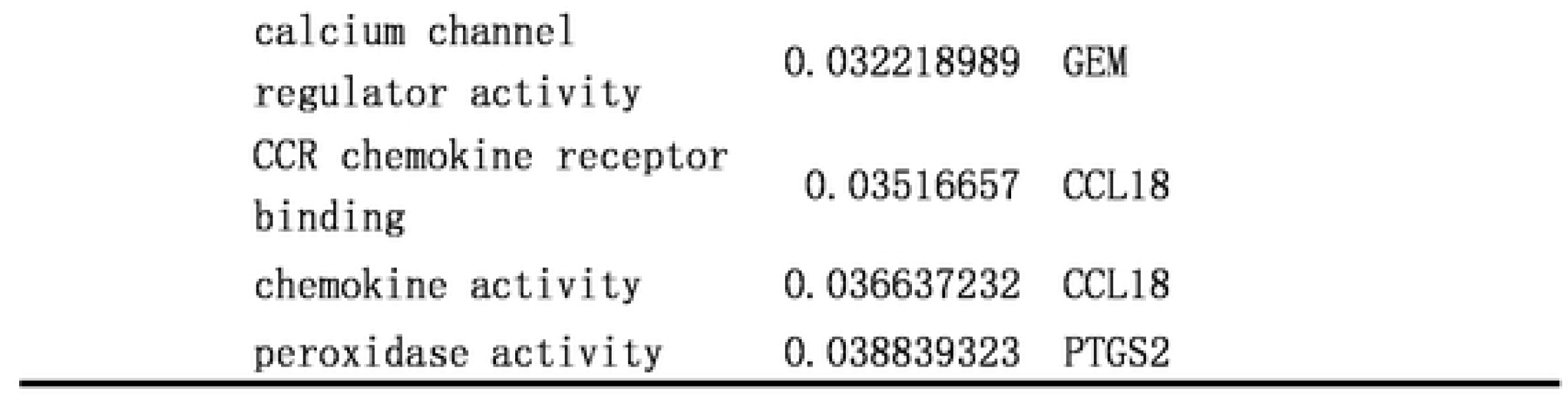
Thctop20MFresultsofGO analysis.

| ID | Description | pvalue | geneID |
| --- | --- | --- | --- |
|  | extracellular matrix structural constituent | 6.85E-06 | CHI3L1/THBS2/CILP/ASPN |
|  | collagen binding | 0.001188741 | MMP9/ASPN |
|  | integrin binding | 0.006072574 | THY1/FAP |
|  | serine-type endopeptidase activity | 0.007501046 | MMP9/FAP |
|  | dipeptidyl-peptidase activity | 0.007580439 | FAP |
|  | serine-type peptidase activity | 0.008976615 | MMP9/FAP |
|  | serine hydrolase activity | 0.009341335 | MMP9/FAP |
|  | arylsulfatase activity | 0.010597647 | SULF1 |
|  | metalloendopeptidase inhibitor activity | 0.012103054 | SPOCK1 |
|  | sulfuric ester hydrolase activity | 0.01285496 | SULF1 |
|  | glycosaminoglycan binding | 0.013237474 | SULF1/THBS2 |
|  | extracellular matrix structural constituent conferring compression resistance | 0.016606525 | ASPN |
|  | oxidoreductase activity, acting on single donors with incorporation of molecular oxygen, incorporation of two atoms of oxygen | 0.01810344 | PTGS2 |
|  | oxidoreductase activity, acting on single donors with incorporation of molecular oxygen | 0.018851105 | PTGS2 |
|  | glycolipid binding | 0.021836478 | THY1 |
|  | Wnt-protein binding | 0.022581502 | SFRP4 |

|  |  |  |
| --- | --- | --- |
| calcium channel<br>regulator activity | 0.032218989 | GEM |
| CCR chemokine receptor<br>binding | 0.03516657 | CCL18 |
| chemokine activity | 0.036637232 | CCL18 |
| peroxidase activity | 0.038839323 | PTGS2 |

**Table 4.** ThcresultsofKEGGanalysis.

| ID | Description | pvalue | geneID |
| --- | --- | --- | --- |
|  | IL-17 signaling pathway | 0.001896872 | PTGS2/MMP9 |
|  | TNF signaling pathway | 0.002681694 | PTGS2/MMP9 |
|  | Leukocyte transendothelial<br>migration | 0.002776944 | MMP9/THY1 |
|  | MicroRNAs in cancer | 0.019359176 | PTGS2/MMP9 |
|  | Human papillomavirus infection | 0.021922529 | PTGS2/THBS2 |
|  | Bladder cancer | 0.029668581 | MMP9 |
|  | Malaria | 0.036082009 | THBS2 |
|  | Ovarian steroidogenesis | 0.036792427 | PTGS2 |
|  | Regulation of lipolysis in<br>adipocytes | 0.041753142 | PTGS2 |
|  | VEGF signaling pathway | 0.042460074 | PTGS2 |
|  | Arachidonic acid metabolism | 0.043872635 | PTGS2 |
|  | Chemical carcinogenesis - DNA<br>adducts | 0.049505527 | PTGS2 |

3.6 Establishment of ceRNA network related to the prognosis of gastric cancer.

Survival data extracted from the TCGA database were combined with ceRNA data to obtain the ceRNA network related to the prognosis of gastric cancer. hsa_circ_0055521/hsa-miR-204-5p/FAP, (hsa_circ_0005051, hsa_circ_0007613, hsa_circ_0045602, hsa_circ_0034398, hsa_circ_0006089) /hsa-miR-32-3p/FNDC1 (Figure 6.1). Then, survival analysis of FAP and FNDC1 genes was performed, P < 0.05 (Figure 6.2).

**Figure 6.1.**
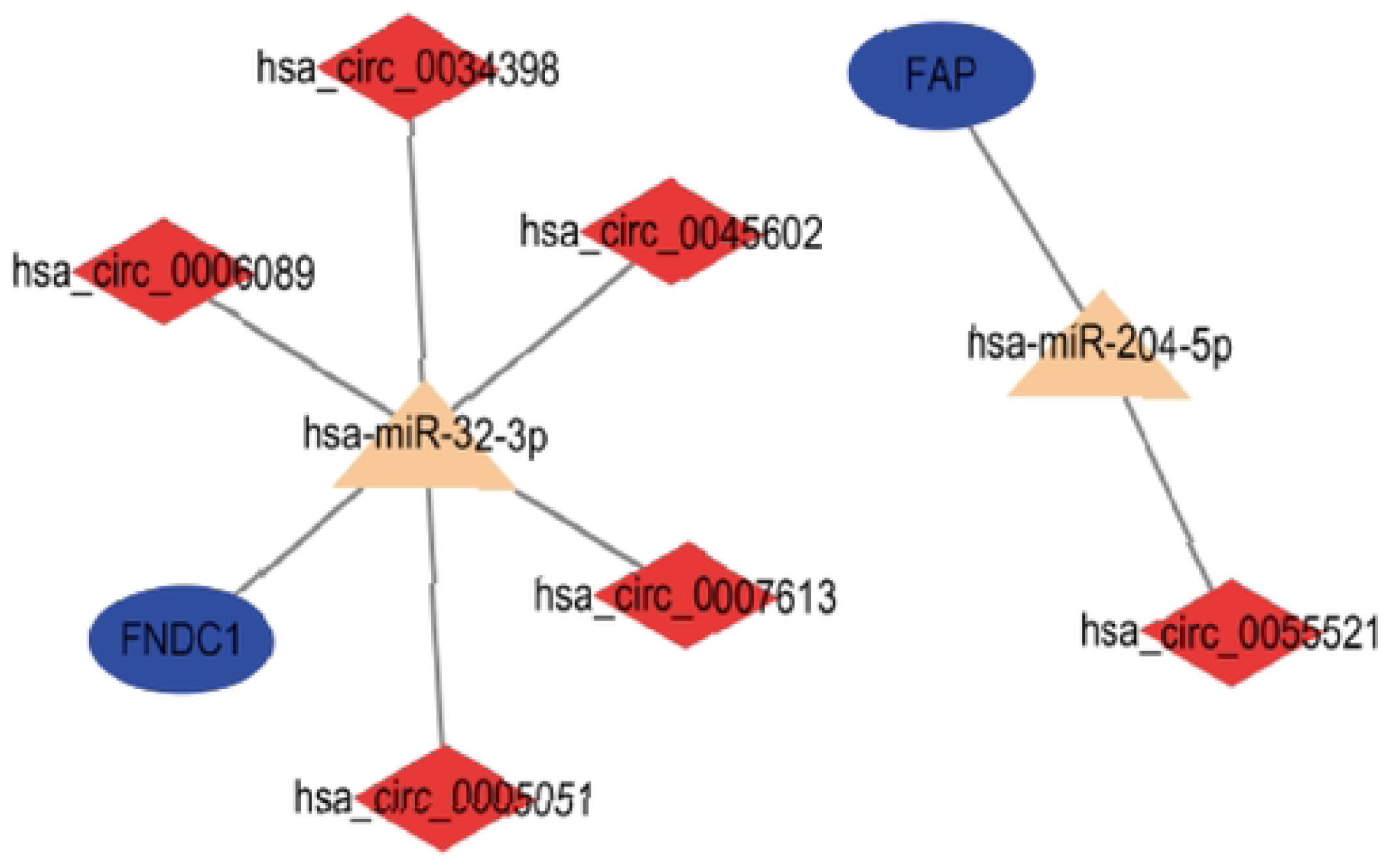
ccRNA networks associated with the prognosis of gastric cancer.

**Figure 6.2.**
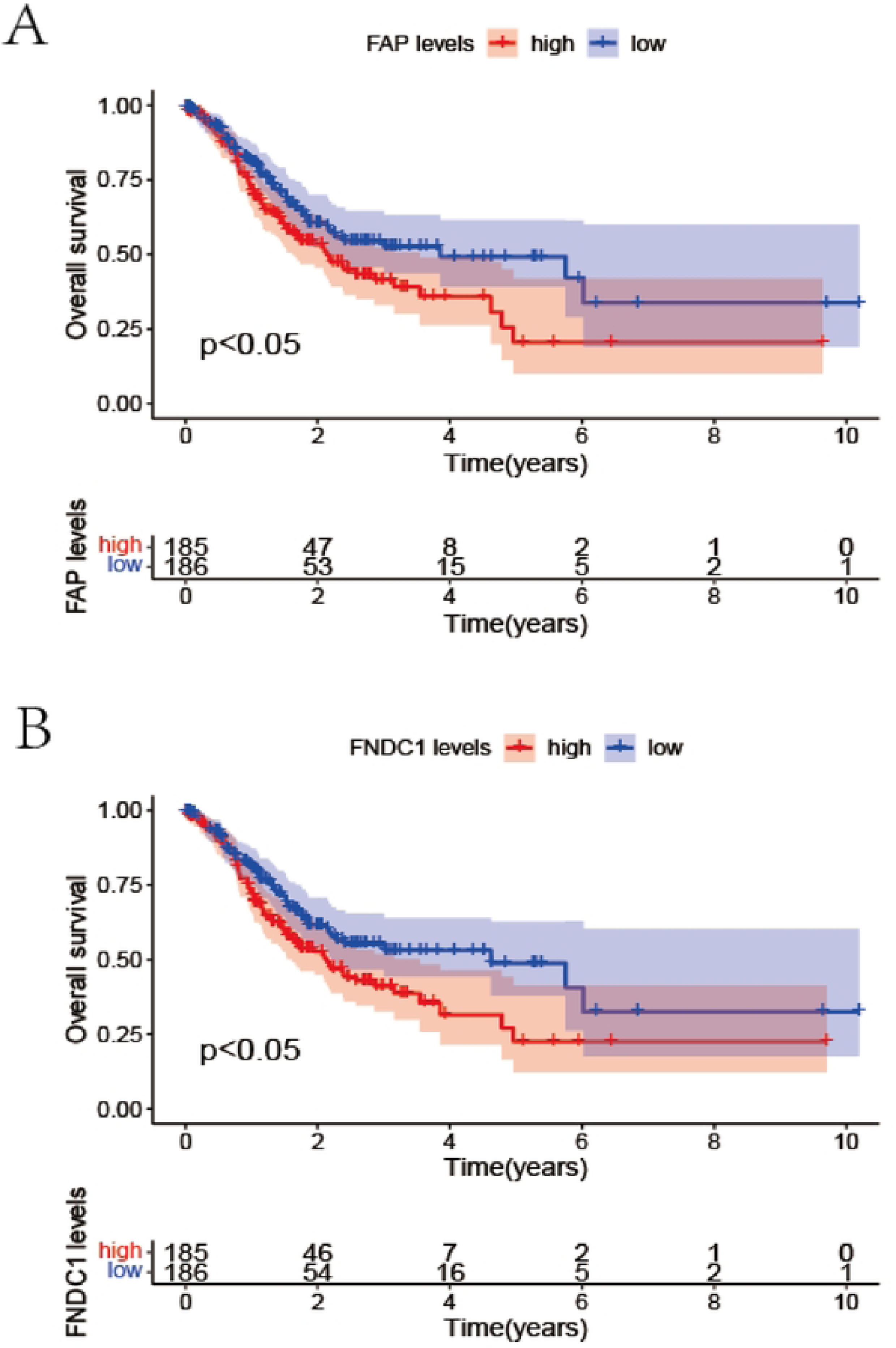
A: Survival analysis of target gene FAP, P < 0.05; B: Survival analysis of target gene FNDCI, P < 0.05.

3.7 Immunohistochemical analysis.

We through the human protein map (the human protein atlas, HPA), org (https://www.proteinatlas. /) network database of FNDC1, FAP this two may affect the prognosis of gastric cancer gene independently, immunohistochemical analysis, The comparison of protein expression of FNDC1 and FAP in gastric cancer tissue and adjacent normal tissue showed significant differences in protein expression of FNDC1 and FAP in gastric cancer tissue and adjacent normal tissue respectively (Figure 7).

**Figure 7.**
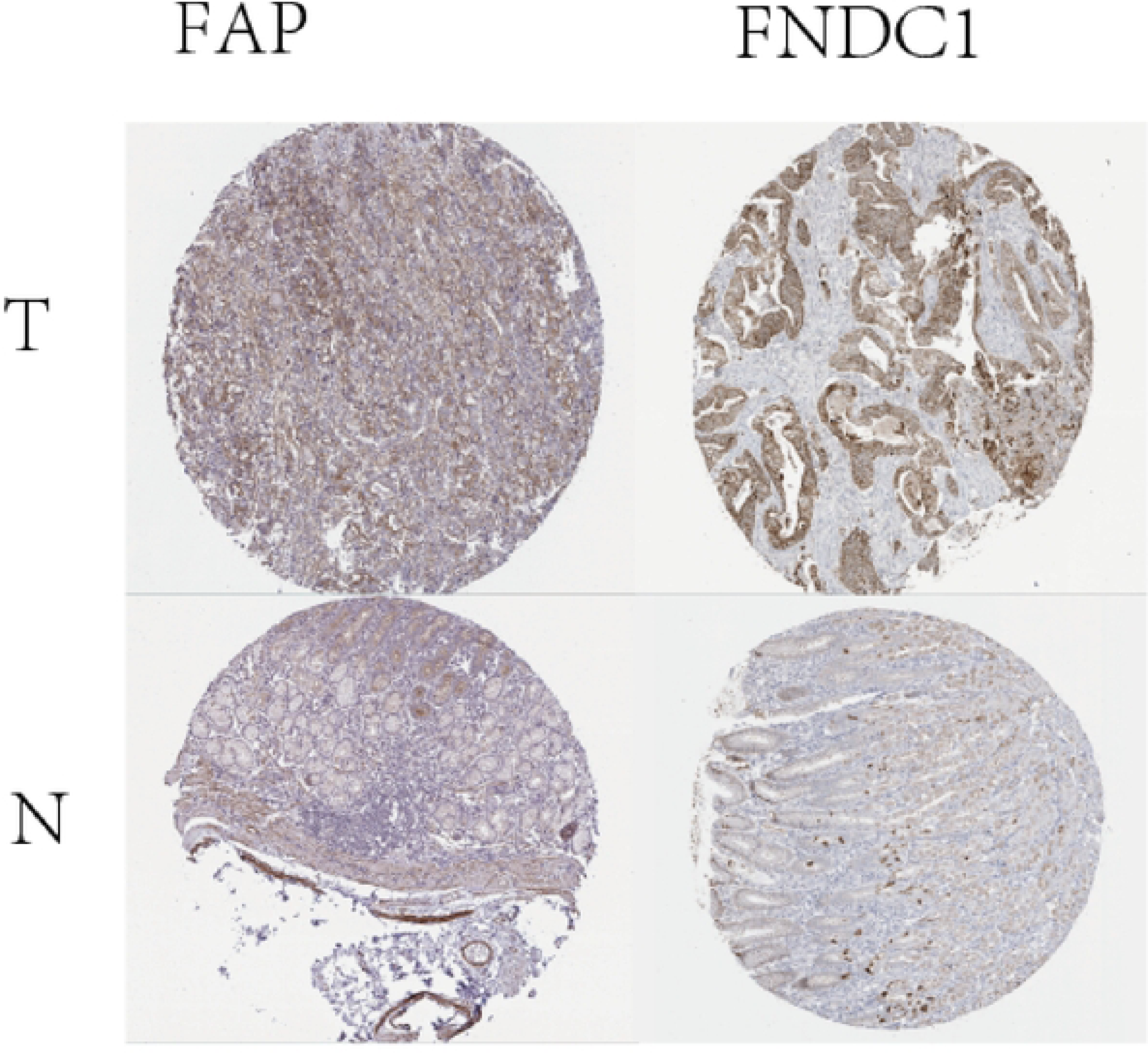
**lmmunohistochemical profiles of target genes FAP and FNOCI based on HPA database.**

## 4. Discussion

At present, non-coding RNA (ncRNA) can be divided into microRNAs (miRNAs), long non-coding RNAs (lncRNAs), circular RNAs (circRNAs) and PIWI interacting RNAs (piRNAs) according to their length, shape and location. As a functional regulatory molecule mediating cellular processes such as chromatin remodeling, transcription, post-transcriptional modification, and signal transduction; The networks in which ncRNAs participate can influence numerous molecular targets to drive specific cellular biological responses and fates, which in turn influence the development and progression of tumors[21,22]. Based on the analysis of gastric cancer data from TCGA and GEO sources, this study explored the ceRNA network composed of circRNAs that may affect the prognosis of gastric cancer and its possible mechanism.

In this study, circRNA, miRNA and mRNA with significant differences in expression between gastric cancer tissues and adjacent normal tissues of gastric cancer were first screened, and the circRNA reaction originals and related mRNA were screened. A new circRNA-based ceRNA network diagram was then constructed. Through GO and KEGG analysis of ceRNA target genes, we found that their main functions were enriched in IL-17 signaling pathway and TNF signaling pathway. Therefore, our study found that these two pathways may be related to the occurrence and development of gastric cancer.

There have been many studies on the role of inflammatory factors in cancer, and this article will not elaborate on them. In terms of IL-17 signaling pathway, this study shows that it may affect the occurrence and development of gastric cancer. Of course, the specific mechanism needs further experimental verification. Previous studies have found that it may promote tumor progression, growth and migration of tumor cells by changing the tumor micro-environment, leading to poor prognosis in patients with gastric cancer[23,24]. Studies of gene network pharmacology have found that IL-17 signaling pathway may be associated with gastric cancer [25]. Previous experimental studies have found that IL-17 may affect the occurrence and development of gastric cancer by regulating Beclin-1 ubiquitination and autophagy[26]. As for the study of TNF signaling pathway, previous studies have found that TNF may affect the micro-environment of gastric carcinoma and the development of gastric cancer[27]. It has also been found that TNF may influence the progression of gastric cancer through signaling mediated epithelial-stromal interaction[28]. These studies further indicate that IL-17 signaling pathway and TNF signaling pathway may affect the development and prognosis of gastric cancer from various aspects.

Two sub-networks related to the prognosis of gastric cancer were screened through prognostic data: hsa_circ_0055521/hsa-miR-204-5p/FAP, (hsa_circ_0005051, hsa_circ_0007613, hsa_circ_0045602, hsa_circ_0034398, hsa_circ_0006089) /hsa-miR-32-3p/FNDC1. Different from previous studies, these two prognostic related ceRNA networks are new circRNA-based prognostic related ceRNA networks for gastric cancer. Through circbase database, we found that the target gene of hsa_circ_0055521 is circ MRPL35, and circ MRPL35 gene has been found in previous studies to inhibit the proliferation of gastric cancer cells [29,30]. The FAP gene identified in this study has no correlation with hsa_circ_0055521 reported in previous studies. However, this study found that hsa_circ_0055521 may regulate the expression of FAP through sponging action on hsa-miR-204-5p, thus regulating the occurrence and development of gastric cancer. It can affect the prognosis of gastric cancer. Through geneCards, we found that FAP is mainly associated with invasive basal cell carcinoma and melanoma. At present, studies have also found that FAP may be related to drug resistance, prognosis and peritoneal metastasis of gastric cancer[31,32]. In the second CircrNa-based gastric cancer prognostic network in this study, it was found that circUBXN7, circPRRC2B, circHN1, circC15orf41 and circASAP2 could regulate the expression of FNDC1 through competitive sponging of hsa-miR-32-3p. It can affect the occurrence, development and prognosis of gastric cancer. Through the screening of previous studies, we found that there are few studies on the effects of circUBXN7 on tumor prognosis, and only the effects of circUBXN7 on the proliferation, migration and inhibition of apoptosis of hepatocellular carcinomates [33]. However, the effect on prognosis of gastric cancer has not been reported. Similarly, for the effect of circPRRC2B in tumors, we found that it may be relevant to the development and survival of patients with nephroblastoma[34]. However, studies on its relationship with gastric cancer and other tumors have not yet found, but this study found that it may be related to the prognosis of gastric cancer. For circHN1, we found that some studies have found an association with the progression of gastric cancer [35-37]. However, there are few studies, which can be used as the direction of further research on the occurrence and development of gastric cancer. Studies on the relationship between circC15orf41 and the occurrence and development of gastric cancer have not yet been found, but we have found through previous studies that it may be related to the metastasis of osteosarcomatus[38]. This also provides a new direction for us to explore the road of gastric cancer treatment. This study found that circASAP2 may be related to the prognosis of gastric cancer, and some previous studies on this aspect have also confirmed that circasap2 may affect the drug resistance and prognosis of gastric cancer [39-42], and of course may also affect the occurrence and development of pancreatic cancer [43]. However, the specific mechanism of its influence on the occurrence, development and prognosis of gastric cancer is not fully understood and needs further exploration. Regarding the second target gene FNDC1 discovered in this study, current studies on its relationship with gastric cancer mainly focus on the possibility that its high expression may be associated with poor prognosis of gastric cancer, but the specific mechanism remains to be further explored [44-48]. Through the above studies, it was found that circUBXN7, circPRRC2B, circHN1, circC15orf41, circASAP2, FNDC1 and FAP may affect the prognosis of gastric cancer, and the specific mechanism needs to be further studied.

Finally, we conducted immunohistochemical analysis of the proteins of the two target genes identified in this study through the HPA database, and we found that there were significant differences in the protein expressions of FNDC1 and FAP in cancer tissues and adjacent normal tissues. These two genes, FNDC1 and FAP, may affect the occurrence and development of gastric cancer and the prognosis of gastric cancer.

## 5. Conclusion

In summary, this study found that IL-17 signaling pathway and TNF signaling pathway may affect the occurrence, development and prognosis of gastric cancer. And hsa_circ_0055521/hsa-miR-204-5p/FAP, (hsa_circ_0005051, hsa_circ_0007613, hsa_circ_0045602, hsa_circ_0034398, hsa_circ_0006089) /hsa-miR-32-3p/FNDC1 two circRNA-based stomachs The cancer ceRNA prognostic network is a new prognostic related ceRNA network for gastric cancer. FNDC1 and FAP may be potential therapeutic targets for gastric cancer.

## ARTICLE HIGHLIGHTS

### Research background

Gastric cancer (GC) is one of the most common malignant tumors, and its pathogenesis are still unclear.

### Research motivation

The present study for the first time investigated the multi-database based ceRNA regulatory network for gastric cancer prognosis using bioinformatics.

### Research objectives

The aims of this study are to explore the multi-database based ceRNA regulatory network for gastric cancer prognosis, so as to provide new strategies for the treatment of GC.

### Research methods

In this study, bioinformatics strategy was used to obtain Datasets from The Cancer Genome Atlas, Gene Expression Omnibus and Gene Expression Profiling Interactive Analysis. The software of R software, circBase and Human Protein Atlas, were performed to analyze and integrate the mRNA datasets, respectively.

### Research results

KEGG analysis of ceRNA’s mRNA showed that the pathway was mainly enriched in IL-17 signaling pathway, TNF signaling pathway and so on, which affected the prognosis of gastric cancer. hsa_circ_0055521/hsa-miR-204-5p/FAP, (hsa_circ_0005051, hsa_circ_0007613, hsa_circ_0045602, hsa_circ_0034398, hsa_circ_0006089) /hsa-miR-32-3p/FNDC1 were the ceRNA networks related to the prognosis of gastric cancer collaterals.

### Research conclusions

This study found that IL-17 signaling pathway and TNF signaling pathway may affect the occurrence, development and prognosis of gastric cancer. And hsa_circ_0055521/hsa-miR-204-5p/FAP, (hsa_circ_0005051, hsa_circ_0007613, hsa_circ_0045602, hsa_circ_0034398, hsa_circ_0006089) /hsa-miR-32-3p/FNDC1 two circRNA-based stomachs cancer ceRNA prognostic network is a new prognostic related ceRNA network for gastric cancer. FNDC1 and FAP may be potential therapeutic targets for gastric cancer.

### Research perspectives

The two circRNA-based stomachs cancer ceRNA prognostic network is a new prognostic related ceRNA network for gastric cancer, which needs further confirmation through molecular biology and clinical experiments.

## Data Availability

All original data files are available from the TCGA,GEO,circBase ect database

## Notes

### Competing Interest Statement

The authors have declared no competing interest.

### Funding Statement

The author(s) received no specific funding for this work

